# The effect of parental enhancing program with mobile application on parental stress and competence among adolescent postpartum women: A quasi-experimental study in Thailand

**DOI:** 10.1101/2025.04.25.25326425

**Authors:** Sunee Kleebpan, Pornpimol Apartsakun, Pensiri Chaiyanusak, Ellen Kitson-Reynolds

## Abstract

**Background:** Adolescent pregnancy constitutes a critical public health issue worldwide. Young mothers face substantial physical and psychological changes as they transition to motherhood, while limited knowledge, experience, and maturity may impair parenting and increase stress. The aim of the study was to evaluate the effects of the parental enhancing program with mobile application on parental stress and competence among adolescent postpartum women.

**Methods:** A quasi-experimental study was conducted among sixty adolescent postpartum women, aged 15-19. They were randomly assigned to receive parental enhancing program with mobile application in intervention group (n = 30) and standard postpartum care in control group (n = 30). Adolescent postpartum women in intervention group were received two sessions in the parental enhancing program before discharge with weekly follow-up through the Line Official Account™ Parent Paplearn. Data was collected using questionnaires, including the Edinburgh Postnatal Depression Scale (EPDS), Parenting Stress Index (PSI-4-SF), and Parenting Sense of Competence Scale (PSOC). Evaluations were conducted during 6-weeks postpartum. Independent t-test was used to compare post-intervention mean scores of parental stress and competence between groups.

**Results:** All enrolled participants completed the study (Intervention: 30; Control: 30). At 6-weeks postpartum, adolescent postpartum women in the intervention group who participated in the parental enhancing program with mobile application through ‘Line Official Account™ Parent Paplearn’ demonstrated significantly lower parental stress (t = 4.50, p < .001) and higher parental competence (t = -4.16, p < .001) compared to the control group.

**Conclusion:** Providing knowledge, skill training, and ongoing support in parenting for adolescent postpartum women through the mobile application significantly reduced parental stress and improved parental competence. Healthcare providers, particularly nurses, are encouraged to integrate such accessible and convenient tools into postpartum care routines. Continuous follow-up post-discharge is recommended to provide ongoing counselling, reduce parental stress, and enhance parental competence in adolescent postpartum women.

## Introduction

Adolescent pregnancy is a major global public health concern, with significant impacts on maternal and child health outcomes. According to the United Nations’ Sustainable Development Goals (SDG) progress report, the global adolescent birth rate declined from 43.3 births per 1,000 women in 2019 to 41.3 in 2023 [1]. In Thailand, the adolescent birth rate declined from 31.3 births per 1,000 women in 2019 to 21.0 in 2022. This decline can be attributed to improved contraceptive support and increased accessibility to comprehensive sexual education and effective contraceptive methods [2]. However, the current rates remain above the 2030 SDG target and Thailand’s goal to reduce births among females aged 15-19 to 25 per 1,000 or lower by 2026 [3].

Adolescent pregnancy remains significant health, social, and economic challenges with widespread implications for adolescent mothers’ well-being [4]. Many of these pregnancies are unplanned, with 69.4 percent of adolescent mothers reporting an absence of readiness for the demands of parenting [5]. Young mothers face physical and psychological changes, a role that imposes new responsibilities and often heightens anxiety and fear related to child-rearing. This transition frequently leads to role conflicts and a struggle to balance their developmental needs with parental duties [6]. Peer influence and societal norms contribute to the perception of adolescent pregnancy as normal, with many teenage mothers feeling prepared and viewing their decisions positively. It has been shown that teenage mothers tend to have a more positive attitude toward pregnancy and childbirth than negative [7]. However, poverty, unemployment, and limited education further complicate their situation [8], while lack of knowledge and maturity increase parenting stress. These challenges highlight the need for support systems and educational resources to enhance parenting confidence and reduce stress [9].

Parental stress encompasses the emotional responses triggered by an individual’s perceptions and interpretations of diverse parenting demands. For adolescent mothers, this new role requires them to learn child-care skills, foster a parent-child relationship, and interpret their child’s behaviors despite limited experience and resources. Many adolescent mothers report stress associated with navigating parenting responsibilities, such as seeking guidance from family and friends, learning through clinical interactions, consulting with experienced peers, and accessing information online [10]. Structured support, including knowledge sharing, skills training, and online platforms facilitated by nurses, can help reduce stress and enhance parenting confidence.

Parental competence reflects a mother’s positive attitude, comprehensive knowledge, and practical skills in child-rearing, enabling effective parenting and fulfilling parenting responsibilities and satisfaction. However, adolescent postpartum women, face significant challenges, leading to increased stress and reduced confidence, which can impair their maternal role [6,11]. Therefore, they require targeted interventions to strengthen parenting skills, bolster their confidence, and enhance their ability to fulfill their role effectively.

In delivering care and promoting the well-being of adolescent postpartum women, nurses play a crucial role by adopting a supportive attitude and fostering open communication to address the unique needs of this population as mentioned in Belsky’s conceptual framework. This approach helps relieve anxiety and supports adolescent mothers in their adjustment to the maternal role [12–13]. Effective care should focus on role adaptation through education, skills training in childcare and breastfeeding, and encouraging mother-infant bonding to enhance maternal confidence. Additionally, providing emotional support and conducting regular mental health assessments are essential for addressing the psychological and emotional dimensions of postpartum adjustment.

With adolescents increasingly relying on mobile applications to seek information, social media has become essential in modern care models [14]. A review of the literature, along with observations from the researcher’s experience in postpartum care, reveals that adolescent mothers often receive care similar to that provided to adult mothers, using lectures and skill demonstrations. Adolescent mothers lack convenient and engaging methods to revisit parenting information, as educational materials are frequently presented in less accessible formats, such as manuals, brochures, or CDs. Furthermore, options for timely consultation are limited, presenting barriers to effective support [15–19]. Research on mobile health (mHealth) interventions, has shown promising outcomes in maternal childcare [20–21]. To address this, the researcher developed a care model using a mobile application through the ‘Line Official Account™ platform’. This platform enables the dissemination of information, images, and videos in a format more appealing to adolescents while offering a private channel for consultation and real-time consultations. A-LINE Brand & Behavior Study 2020 survey on the behavior of LINE application users in Thailand revealed that, out of 69 million internet users, 47 million use Line applications. Among Generation Z users (under 20 years old), social media activities and communication via the Line application account for 96% of their online time [22]. The Line application is a highly relevant platform for youth engagement.

The researchers developed a parental enhancing program with mobile application through ‘Line Official Account™ Parent Paplearn’ to provide adolescent postpartum mothers with personalized education and skills training in childcare. This program covers topics, including recognizing and responding to infant cues, basic infant care, breastfeeding techniques, bathing procedures, and identifying abnormal symptoms in infants, delivered through infographics, e-books, and videos, serving as readily accessible resources for review and offering a reliable platform for ongoing consultation. The program’s objective is to enhance parenting competence and reduce parenting stress, empowering adolescent mothers with the knowledge and confidence to motherhood.

## Research framework

This study applies Belsky’s “A Process Model of Competent Parental Functioning” [23] to elucidate the various factors influencing parental roles in childcare. This model identifies three domains including 1) ‘Personal psychological resources of parents’ domain encompasses individual characteristics. It has been found to exert the most significant influence on parental functioning. 2)‘Contextual sources of stress and support’, which can either facilitate or hinder effective parenting, and 3) ‘Characteristics of the Child’, which may also influence parenting dynamics but generally have a lesser impact compared to the other two domains.

The researchers utilized the parental competence framework to design the parental enhancing program with mobile application through ‘Line Official Account™ Parent Paplearn’. This program specifically targets contextual sources of stress and support, positioning nurses as vital resources to assist adolescent mothers in effectively caring for their children. In Thailand, nurses play a pivotal role in providing care through the nursing process, providing clients with appropriate education, and offering counselling. The program emphasizes the provision of knowledge and training in essential childcare skills, thereby enabling these mothers to perform their parenting roles with increased competence. Furthermore, it offers continuous support via mobile application through the ‘Line Official Account™ platform, facilitating timely responses to questions and concerns. This ongoing support not only alleviates parenting-related stress but also enhances the mothers’ overall competence in childcare (Fig 1).

**Fig 1.**
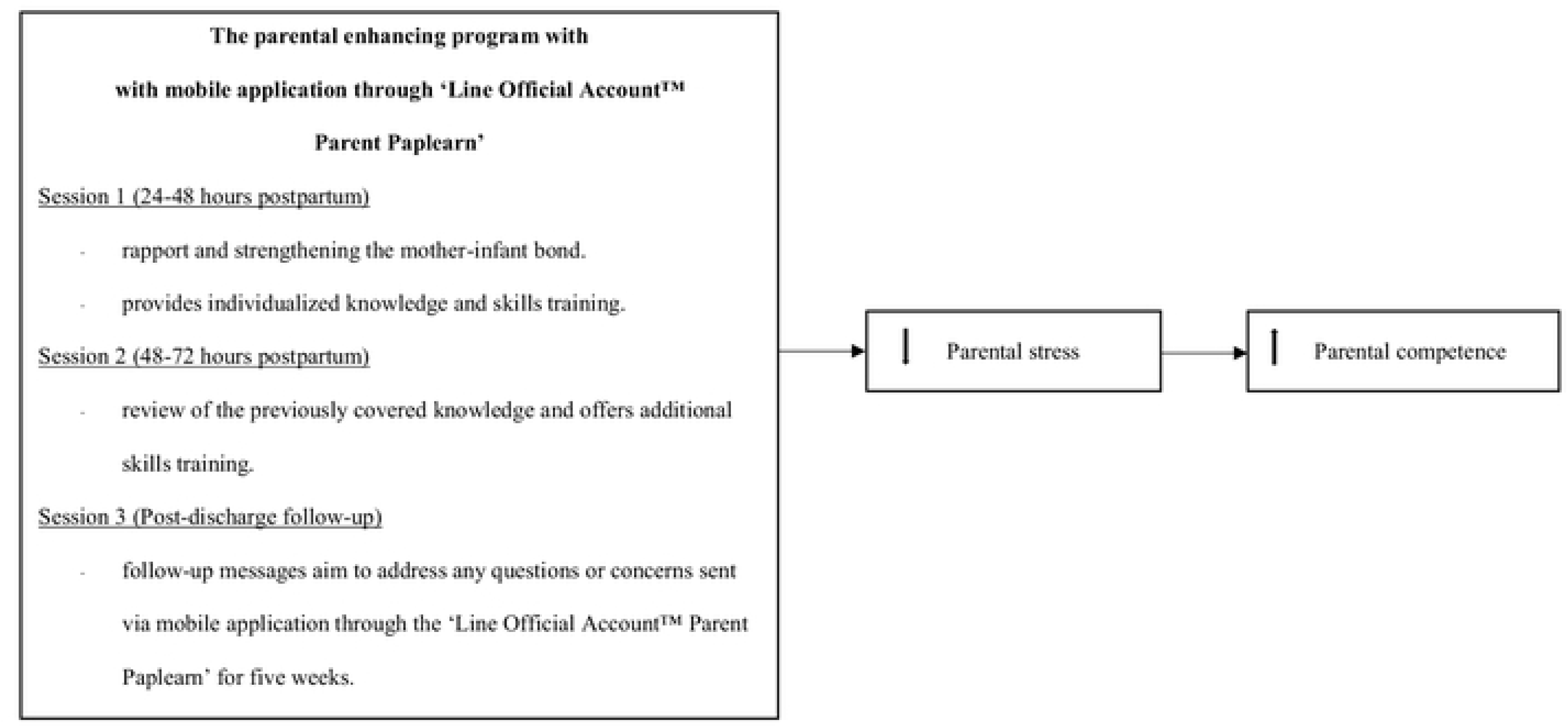
Research framework.

## Materials and methods

### Study design

This study employed a quasi-experimental control group pre-post-test design to compare parental stress and competence between adolescent postpartum mothers who participated in the parental enhancing program with mobile application through ‘Line Official Account™ Parent Paplearn’ and those receiving standard postpartum care.

### Setting and samples

The study was conducted at a tertiary care hospital, Queen Savang Vadhana Memorial Hospital, Thai Red Cross Society, Thailand. The hospital has an average 3,800 of postpartum women per year, with approximately 2.5 percent being adolescent postpartum women. The study sample consisted of postpartum women aged 15 to 19 who were admitted to the general obstetrics ward. All participants had full-term deliveries without postpartum complications for either mother or infant. They intended to childcare for six weeks and demonstrated the ability to communicate via mobile using the LINE application. The sample size for this study was determined using G*Power (version 3.1.9.4), the effect size indicated a large effect (Cohen’s guidelines), a power of test equal to 0.80, and a significance level set at 0.05. The analysis indicated that a sample size of 26 participants per group. To account for dropouts of 10 percent, the final sample was 30 participants per group. Consequently, the total sample size was established at 60 participants.

## Research instruments

### Instruments for implementation

**The parental enhancing program**, developed by the researcher, is based on Belsky’s “A Process Model of Competent Parental Functioning” [23]. The program consists of the following activities:

Session 1 (24-48 hours postpartum): Establishes rapport and strengthens the mother-infant bond. The researcher provides personalized knowledge and skills training on recognizing infant cues and responsiveness, general infant care, breastfeeding techniques, bathing procedures, and identifying abnormal symptoms in infants, lasting about 60 to 90 minutes. Teaching materials, such as infographics, e-books, and instructional videos, are disseminated through the ‘Line Official Account™ Parent Paplearn’ as a mobile application.

Session 2 (48-72 hours postpartum): Review the previously covered knowledge and offer additional skills training aimed at enhancing the mother’s confidence. Individualized support is provided to address her specific concerns, lasting about 60 minutes.

Session 3 (Post-discharge follow-up): Following discharge, the program includes weekly follow-up messages sent via mobile application through the ‘Line Official Account™ Parent Paplearn’ for five weeks to address any questions or concerns related to challenges faced by the mother or her infant.

**The Line Official Account™ Parent Paplearn** is a mobile application that facilitates the exchange of text messages, images, and videos on a broad scale. This platform also provides a private channel for adolescent postpartum women, enabling them to send messages for consultations or discussions regarding their parenting concerns. The platform is an online tool designed to enable users to create accounts, customize application designs, manage content, and perform system administration functions. The content available on ‘Line Official Account™ Parent Paplearn’ encompasses essential topics such as recognizing infant cues and responsiveness, general infant care, breastfeeding practices, bathing techniques, and identifying abnormal symptoms in infants. This information is delivered through engaging formats, including infographics, e-books, and videos, designed to enhance understanding and retention among adolescent mothers. The researchers developed the application interface, developed content, and primarily managed the system to address questions from adolescent postpartum women. Participants can review the knowledge as needed, without any limitation on the number of times.

### Instruments for data collection

**The demographic characteristics questionnaire** collects data on the mother’s age, marital status, education level, occupation, household income (THB), sufficiency of income, family structure, desire for children, plan for future education, childcare support, experience in childcare, number of pregnancies, type of delivery, and the infant’s APGAR score.

**The postpartum problem and advice record** was developed by the researcher to systematically document follow-up interactions via mobile application through the ‘Line Official Account™ Parent Paplearn’ after hospital discharge, recording the date, problem, and advice provided.

**The Edinburgh Postnatal Depression Scale (EPDS)** was translated into Thai by Vacharaporn et al. [24] This screening tool consists of 10 items. For example, I have been able to laugh and see the funny side of things, I have looked forward with enjoyment to things, etc. The total scores range from 0 to 30 points. A cut-off score of 11 or higher indicates potential depression. The EPDS serves as an important instrument in assessing the mental health needs of this population, facilitating timely interventions to promote overall well-being.

**The Parenting Stress Index Fourth Edition Short Form (PSI-4-SF)** utilized in this study employed the Thai version developed by Srikosa et al. [25] Originally 36 items, it was reduced to 15 to align with the context of the research, specifically focusing on infants aged six weeks postpartum. It measures three domains including 1) Parental distress, 2) Parent-child dysfunctional interaction, and 3) Difficult child, utilizing a five-point Likert scale. Scores range from 15 to 75 points, where lower scores indicate lower levels of parenting stress, while higher scores reflect greater parenting stress. The instrument demonstrated strong validity (CVI = 0.94) and reliability (Cronbach’s alpha = 0.78).

**The Parenting Sense of Competence Scale (PSOC)** was translated into Thai by Kleebpan et al. [26] This scale consists of 16 items measuring parenting efficacy and satisfaction. For example, the problems of taking care of a child are easy to solve once you know how your actions affect your child, an understanding you have acquired, etc. Scores ranged from 16 to 96 points. In this context, lower scores indicate lower levels of parenting competence, while higher scores reflect greater competence in parenting. The instrument demonstrated strong validity (CVI = 0.81) and reliability (Cronbach’s alpha = 0.72).

### Ethical considerations

This study received ethical approval from the Ethics Committee of Queen Savang Vadhana Memorial Hospital, Thai Red Cross Society (COE No. 001/2566) on February 6, 2023, the study has been registered on Thai clinical trials registry (TCTR20250309010, https://www.thaiclinicaltrials.org/show/TCTR20250309010). Participants were informed about the study, assured of confidentiality, and provided written consent, with parental or spousal consent required for those under 18. Participation was voluntary, with the right to withdraw at any time without any consequence. Participants scoring 11 or higher on the EPDS or indicating suicidal thoughts were not excluded. Instead, the researcher provided initial support by actively listening to their concerns and referred them to a nurse who facilitated further consultation with a psychiatrist, ensuring that they received appropriate emotional and mental health care.

### Data collection procedure

The study participants were recruited from 06 February 2023 to 21 June 2024. The researcher collaborated with nurses to identify eligible participants who met inclusion criteria and obtained permission to meet them after 24 hours postpartum. During these meetings, the researcher explained the study objectives, procedures, and participants’ rights. Informed consent forms were then obtained from the participants. Before the intervention, the researcher administered several assessment tools to collect baseline data, including the demographic characteristics questionnaire, the Edinburgh Postnatal Depression Scale [24], the Parenting Stress Index Fourth Edition Short Form [25], and the Parenting Sense of Competence Scale [26]. After that, all participants were scheduled for intervention, while the control group had standard postpartum care. This process will continue until the required sample size is reached, with 30 participants in each group. The research will be conducted in the control group first, followed by the intervention group, to prevent confounding factors.

At one week postpartum, participants in both the intervention and control groups completed the Edinburgh Postnatal Depression Scale [24] to screen for depression, ensuring that appropriate emotional and psychological support could be provided based on the findings. Then, the researcher sent weekly messages via a mobile application through the Line Official Account™ Parent Paplearn for a duration of five weeks. These messages were designed to address any questions or concerns regarding the challenges participants faced in child-rearing and parenting responsibilities after discharge, based on postpartum problem and advice records. At the conclusion of the study, six weeks postpartum, participants in both groups were administered the Parenting Stress Index Fourth Edition Short Form [25] and the Parenting Sense of Competence Scale [26]. Details of the study as shown in Fig 2.

**Fig 2.**
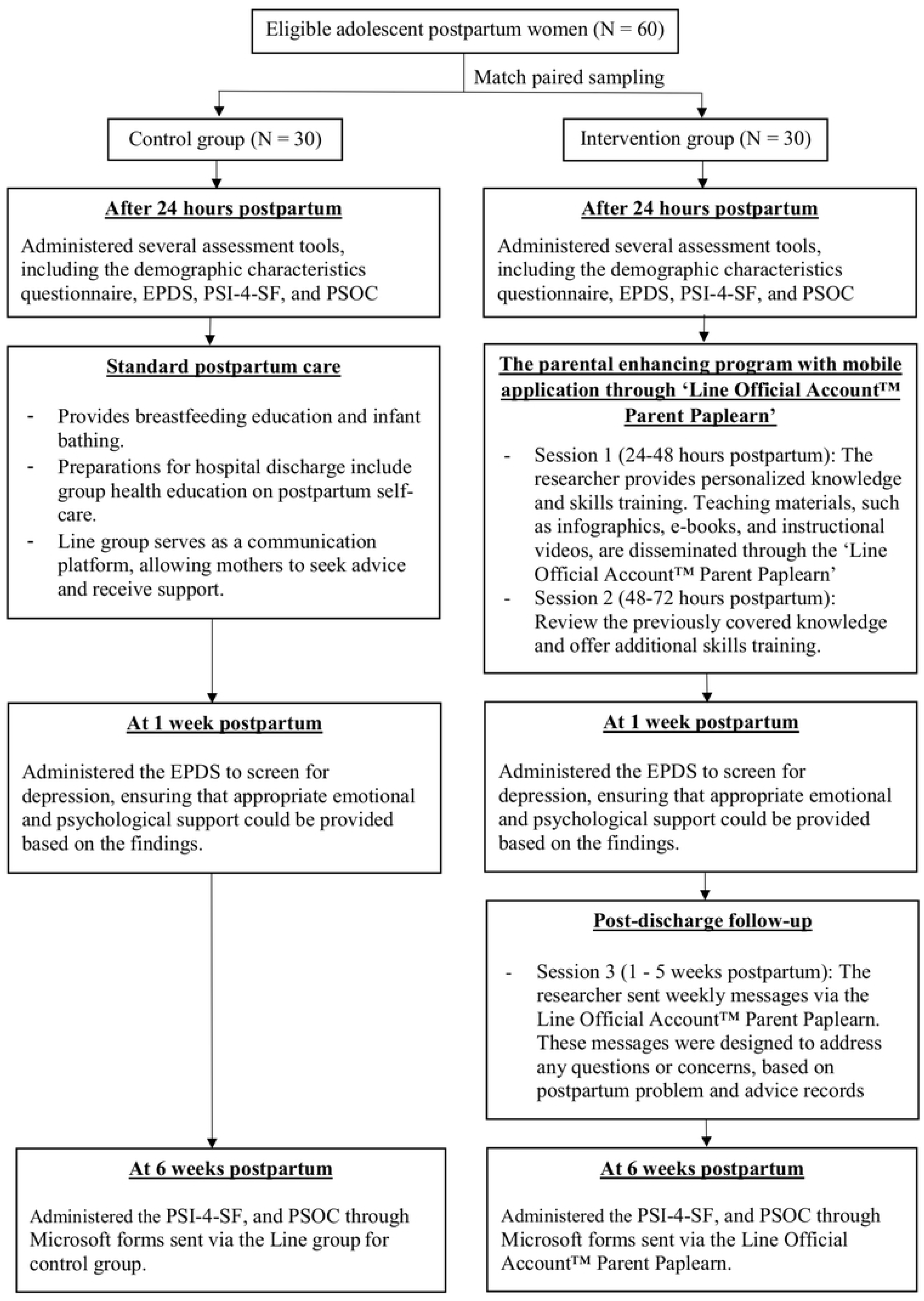
Flow diagram of study.

### Statistical analysis

IBM SPSS Statistics Version 28 (License code: 5f5119869c4164e0bda0) was used for data analysis. Descriptive statistics were used to characterize the participant demographic and obstetric characteristics. The comparison of differences between the characteristics of the intervention group and the control group was conducted using independent t-test, Chi-square test, and Fisher’s exact test, based on type of the variables and assumptions of statistical. The independent t-test was used to determine significant differences in parental stress and competence scores between the intervention group and the control group with statistical significance was set at p < .05. Prior to data analysis, it was checked to ensure that the data met the statistical assumptions.

## Results

In the study 30 adolescent postpartum women in each group, there were founded that no significant differences between the intervention and control groups in all demographic and obstetric characteristics, except for the significant difference in EPDS at 1 week postpartum. The details are shown in Table 1.

**Table 1.**
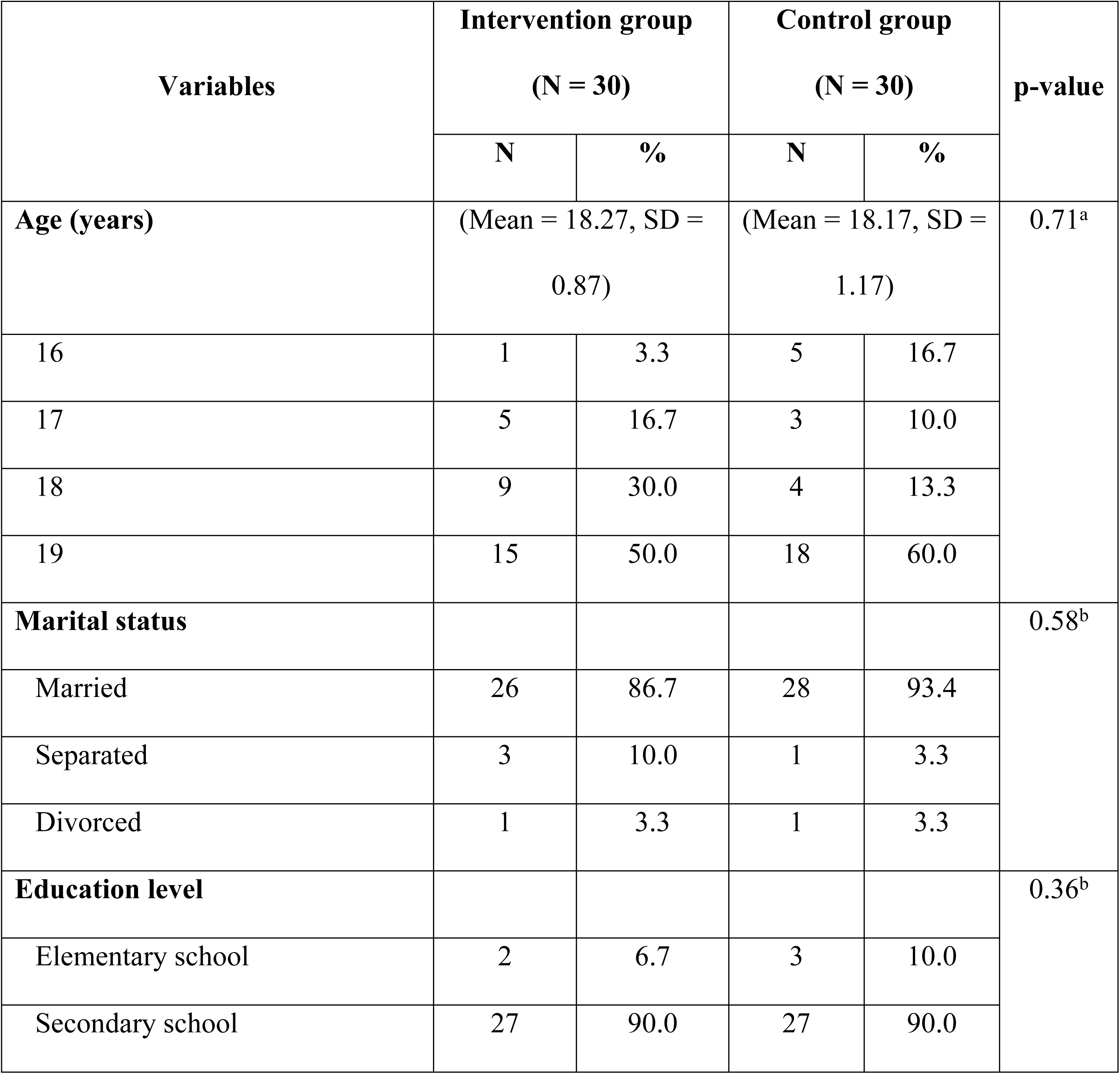
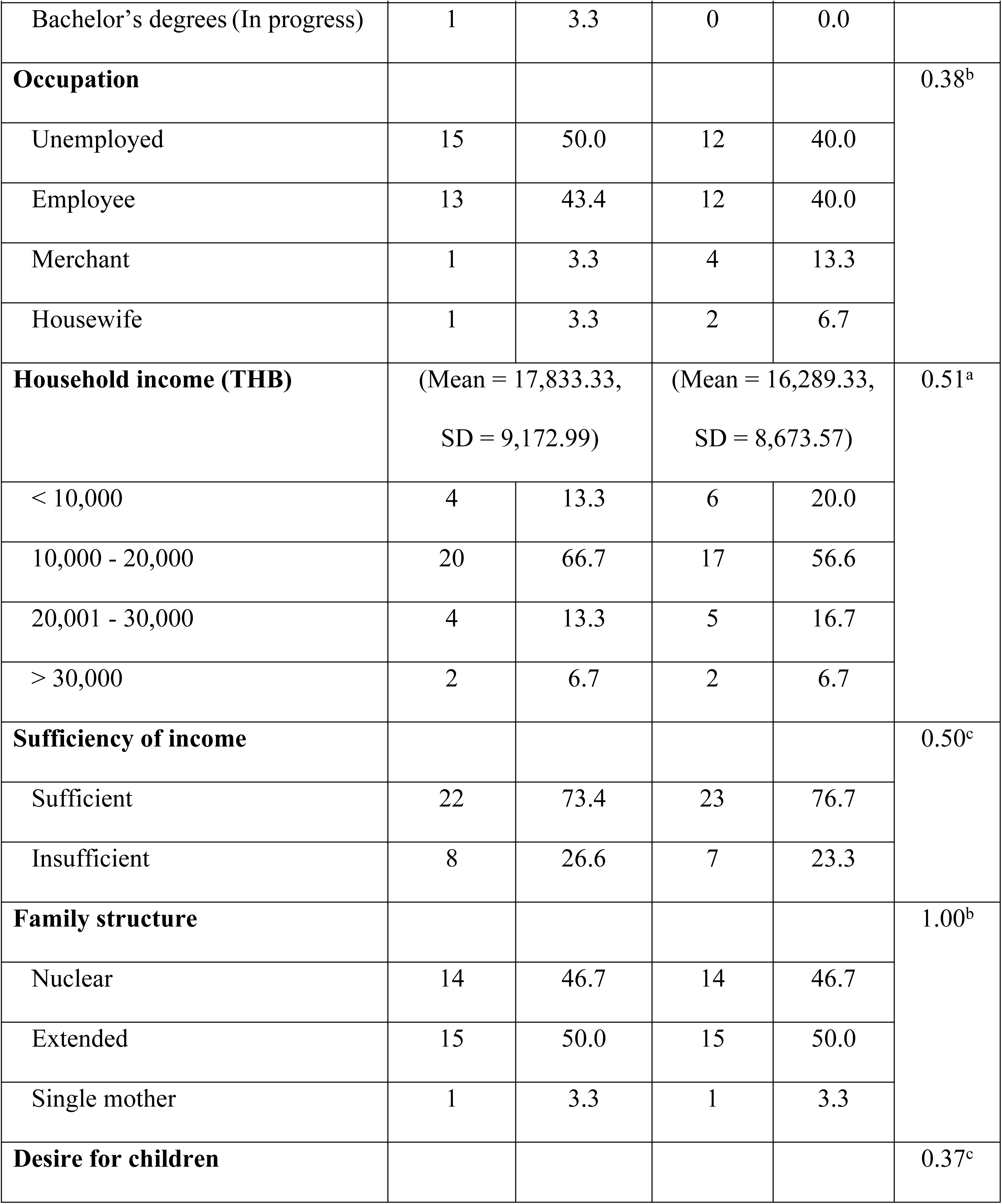
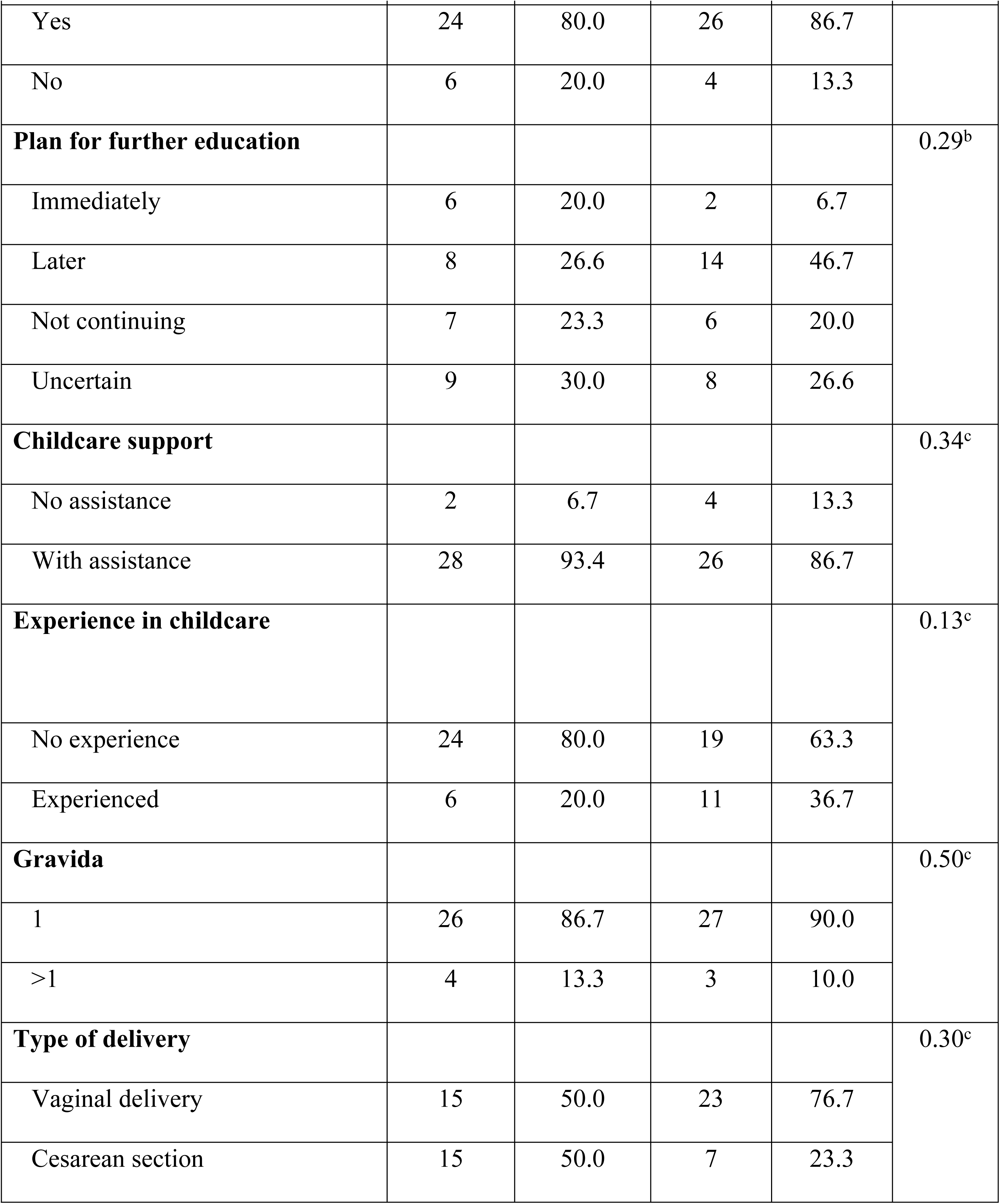
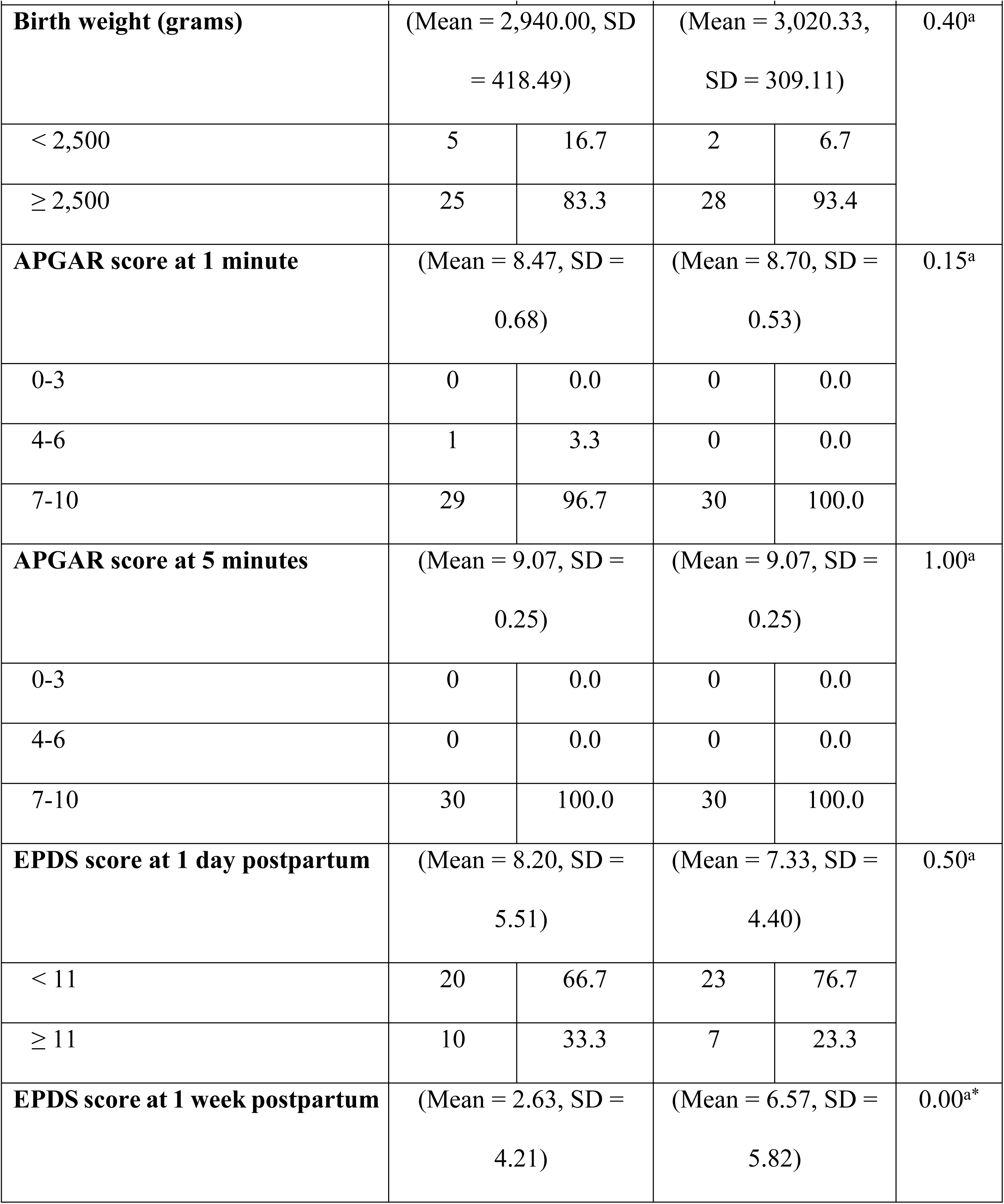
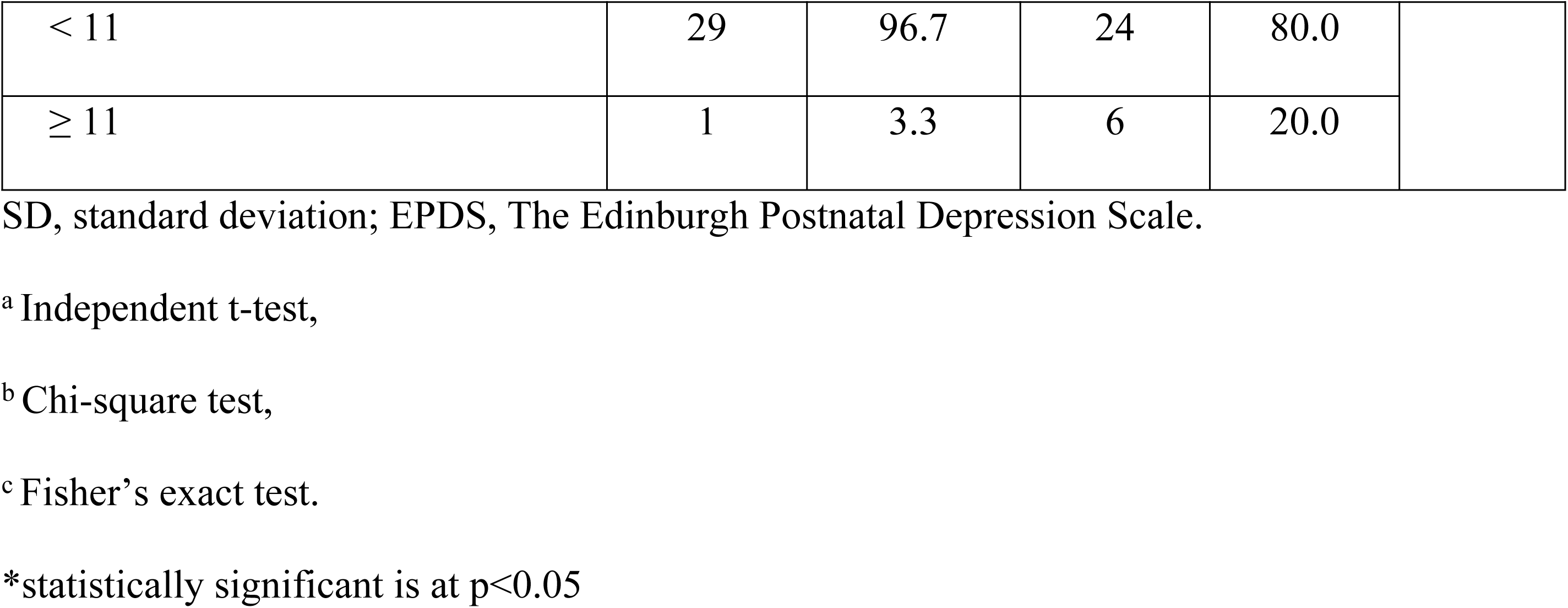
Participant demographic and obstetric characteristics.

The paired t-test revealed a statistically significant difference in the intervention group after the parental enhancing program with mobile application through ‘Line Official Account™ Parent Paplearn’. Parental stress significantly decreased (p < 0.05), while parental competence increased (p < 0.001). In contrast, the control group showed no statistically significant differences in either parental stress (p > .05) or competence (p > .05), as detailed in Table 2.

**Table 2.** Mean scores of parental stress and parental competence before and after the intervention within each group.

| Variables | Before<br>intervention |  | After<br>intervention |  | Mean<br>difference | t-test | p-value |
| --- | --- | --- | --- | --- | --- | --- | --- |
|  | Mean | SD | Mean | SD |  |  |  |
| **Parental stress** |  |  |  |  |  |  |  |
| Intervention group (N = 30) | 37.63 | 7.10 | 32.47 | 4.88 | 5.17 | 3.46 | **0.002\*** |
| Control group (N = 30) | 39.27 | 7.74 | 42.63 | 11.67 | -3.37 | -1.67 | 0.107 |
| **Parental competency** |  |  |  |  |  |  |  |
| Intervention group (N = 30) | 62.77 | 8.45 | 70.87 | 6.49 | -8.10 | -4.55 | **0.000\*** |
| Control group (N = 30) | 63.90 | 6.74 | 62.57 | 8.79 | 1.33 | 0.87 | 0.393 |

SD, standard deviation
\*statistically significant is at $p < 0.05$

At 6-weeks postpartum, the intervention group had lower mean scores of parental stress (Mean = 32.47, SD = 4.87) than the control group (Mean = 42.63, SD = 11.37), with a statistically significant difference (t = 4.50, p < .001). Additionally, mean scores of parental competence was higher in the intervention group (Mean = 70.87, SD = 6.49) compared to the control group (Mean = 62.57, SD = 8.79), with a statistically significant difference (t = -4.16, p < .001), as detailed in Table 3.

**Table 3.** Mean scores of parental stress and parental competence between intervention groups and control groups after intervention (6 weeks postpartum).

| Variables | Intervention group<br>(N = 30) |  | Control group<br>(N = 30) |  | Mean difference | t-test | p-value |
| --- | --- | --- | --- | --- | --- | --- | --- |
|  | Mean | SD | Mean | SD |  |  |  |
| Parental stress | 32.47 | 4.87 | 42.63 | 11.37 | 10.17 | 4.50 | **0.000\*** |
| Parental competency | 70.87 | 6.49 | 62.57 | 8.79 | -8.30 | -4.16 | **0.000\*** |

SD, standard deviation
\*statistically significant is at $p < 0.05$

## Discussion

The findings of the study indicated that participants in the intervention group had significantly lower parental stress and higher parental competence than the control group. These results provide strong support for the research hypothesis.

The parental enhancing program with mobile application through ‘Line Official Account™ Parent Paplearn’, was developed based on Belsky’s “A Process Model of Competent Parental Functioning” [23]. This program specifically addresses contextual sources of stress and support, positioning nurses as essential facilitators in helping adolescent mothers by providing knowledge and skill training in childcare, the program empowers these young mothers to perform their parenting roles with greater competence. Furthermore, it provides ongoing support through the Line platform. The program’s positive impact on reducing parental stress and enhancing parental competence aligns with previous research. For instance, the study by Léniz-Maturana et al [27] demonstrated that maternal role confidence is associated with reduced stress levels. Similarly, Flaherty et al [28] found that parental competence significantly influences parental stress, while perceived social support plays a critical role in moderating stress levels. Overall, the findings of the current study underscore the effectiveness of the parental enhancing program in supporting adolescent mothers during their transition into parenthood.

The findings of this study are consistent with previous research demonstrating the effectiveness of parental programs in reducing stress and enhancing maternal outcomes. Saleh et al [19] found that parenting coaching interventions for teenage mothers significantly reductions in parenting stress, and improvements knowledge, attitudes, behaviors, self-efficacy, and overall maternal functioning. Kordi et al [16] highlighted the importance of maternal role training in achieving maternal role fulfilment and satisfaction. Similarly, Moudi et al [17] showed that brief face-to-face parenting skills training significantly enhanced perceived parenting self-efficacy and mother-infant bonding. These findings highlight the importance of structured support and resources, the program addresses the unique challenges faced by adolescent mothers, thereby fostering their confidence and competence in their new roles. The findings of Polgaya et al [18] underscore the critical importance of continuous support, which includes repeated skills training, regular assessments, and follow-up consultations at four weeks postpartum. Their research highlights how such sustained assistance can significantly enhance maternal attitudes and improve maternal role performance among adolescent mothers. The parental enhancing program with mobile application through ‘Line Official Account™ Parent Paplearn’ embodies this model of ongoing support, the program facilitates an environment where they can ask questions, seek advice, and address their concerns as they navigate the complexities of their new parenting roles.

This study builds upon previous research by integrating a mobile application through ‘Line Official Account™ Parent Paplearn’ to improve accessibility and engagement for adolescent mothers. Shorey et al [21] demonstrated that technology-based supportive educational parenting programs, such as telephone-based interventions and mobile health app follow-ups substantially enhance parenting self-efficacy, strengthen parental bonding, increase perceived social support, and improve overall parenting satisfaction, while also reducing instances of postnatal depression. Chua et al [20] indicated that mobile applications improved parenting outcomes such as parenting self-efficacy, anxiety, depression, social support, and parent-child bonding. Additionally, the findings from the study by Nuampa et al [29] indicated that adolescent mothers perceive mobile applications as valuable and easily accessible sources of information. Furthermore, professional nurses acknowledge that these applications can reduce their workload, improve preparedness, and enhance the standard of support provided. Therefore, the parental enhancing program with mobile application through ‘Line Official Account™ Parent Paplearn’ is highly recommended for adolescent postpartum mothers, as it reduces parental stress, and improves parental competence. The integration of mobile application in this program further ensures accessibility and engagement, making it a valuable resource for adolescent mothers navigating the challenges of early motherhood.

## Study limitations

The researcher developed a parental enhancing program with mobile application through ‘Line Official Account™ to reduce parental stress and improve parental competence. However, effective counselling requires sufficient time from nurses. Additionally, challenges may exist for mothers who lack access to smartphones or are unfamiliar with using the Line application, potentially limiting their participation in the program.

## Conclusion

The parental enhancing program with mobile application through ‘Line Official Account™ effectively reduces parental stress and enhances competence in postpartum adolescent mothers. Nurses should advocate for the use of the mobile application for accessible parenting support. Continuous follow-up post-discharge is essential for counselling, alleviating parental stress, and improving parental competence. If this program is not successfully implemented, adolescent mothers may miss crucial opportunities for accessible information and timely counseling. Consequently, this may result in increased parenting stress, decreased confidence in their parenting abilities, and delayed parenting competence. Future research should explore alternative technologies to enhance support.

## Data Availability

All relevant data are within the manuscript.

## Acknowledgement

The authors would like to thank all the participants who contributed to this study.

## Author contributions

**Conceptualization:** Sunee Kleebpan

**Data curation:** Sunee Kleebpan, Pensiri Chaiyanusak

**Formal analysis:** Sunee Kleebpan, Pornpimol Apartsakun

**Funding acquisition:** Sunee Kleebpan

**Investigation:** Sunee Kleebpan, Pornpimol Apartsakun, Pensiri Chaiyanusak

**Methodology:** Sunee Kleebpan, Pornpimol Apartsakun

**Project administration:** Sunee Kleebpan

**Resources:** Sunee Kleebpan

**Supervision:** Sunee Kleebpan, Pornpimol Apartsakun

**Validation:** Sunee Kleebpan, Pornpimol Apartsakun, Pensiri Chaiyanusak, Ellen Kitson-Reynolds

**Visualization:** Sunee Kleebpan

**Writing-original draft:** Sunee Kleebpan, Pornpimol Apartsakun, Ellen Kitson-Reynolds

**Writing-review & editing:** Sunee Kleebpan, Pornpimol Apartsakun, Ellen Kitson-Reynolds

